# S4-Oligo: optimizing polygenic score methods for clinical deployment

**DOI:** 10.64898/2026.10.16.26365773

**Authors:** John Baierl, Jonathan P. Tyrer, Daniel R. Barnes, Michelle Jones, Pei-Chen Peng, Marc Tischkowitz, Antonis C. Antoniou, Paul D. P. Pharoah

**Author notes:** Corresponding author: Paul D. P. Pharoah. These authors contributed equally to this work (John Baierl, Jonathan P. Tyrer).

## Abstract

Polygenic scores (PGSs) have progressed toward clinical utility in recent years, demonstrating odds ratios that are comparable with both lifestyle-related and monogenic risk factors for several complex diseases. However, PGS implementation relies on developing analytically and clinically valid assays. Methods for developing polygenic models (PGMs) that incorporate tens of thousands to millions of variants require either genome-wide chip genotyping combined with imputation or whole genome sequencing, limiting their portability. Additionally, variants may be in regions where the local genomic sequence makes it difficult to assay, and multi-allelic or strand ambiguous variants can make bioinformatics more complicated. Because of these challenges, attempts to develop a reliable assay for published PGMs can produce multiple variations from the same underlying model, hindering routine clinical deployment and incorporation into multi-factorial risk prediction models. In this analysis, we present an adjustment to the previously published select and shrink with summary statistics (S4) PGM method designed to optimize performance for realistic clinical use by restricting to a pre-specified number of available SNPs that are well-captured by genotyping arrays. We compare the performance of this oligogenic S4 method (S4-Oligo) to currently published PGSs as well as the full S4 model for prediction of breast, colorectal, ovarian, and prostate cancers in the *All of Us* Research Program, Biobank Japan, and UK Biobank. We find that S4-Oligo matches or exceeds the accuracy of published PGMs at comparable model sizes while retaining between 88% and 99% of the performance of the original S4 model when limited to 1,000 variants.

## INTRODUCTION

Polygenic scores (PGSs) have progressed toward clinical utility in recent years, demonstrating odds ratios that are comparable with both lifestyle-related and monogenic risk factors for several complex diseases [1,2]. By capturing the additive risk component of common germline genetic variation, PGSs contribute information that improves the accuracy of prediction models independent of traditional risk factors and offer benefits that underlie their potential for clinical use [3]. Since PGSs measure germline variation, they only need to be measured once in an individual’s lifetime and capture aspects of disease risk that precede traditional risk factors. This can refine early prevention strategies and targeted screening through incorporation into multi- factorial risk prediction models. For example, PGSs are currently included in the CanRisk comprehensive risk tool for breast, ovarian, and prostate cancer risk estimation [4,5].

Genome-wide association studies (GWAS) were designed to detect associations with individual single nucleotide polymorphisms (SNPs) meeting strict statistical significance thresholds to limit type I error. However, prediction with PGSs allows for a more lenient signal-to-noise tradeoff to optimize performance at the population level. Including a large number of SNPs in the PGM rather than restricting to those reaching genome-wide statistical significance has been shown to substantially increase predictive power [6–8]. As a result, methodological advances have led to the development of PGM methods that incorporate tens of thousands to millions of variants [9–11]. Computing PGSs based on these highly polygenic models requires either genome-wide chip genotyping data combined with imputation, or whole genome sequencing.

While this has meaningfully advanced research in disease risk prediction, PGS implementation relies on developing analytically and clinically valid assays. Clinical genetics laboratories often combine panel gene sequencing with sequencing of sets of SNPs to generate a PGS. This method is generally suitable for hundreds of SNPs. However, even for published PGMs with fewer than 1,000 SNPs, such as the PGS313 for breast cancer, developing a sequencing-based assay is non-trivial for several reasons [7]. Variants may be located in regions where the local genomic sequence makes it difficult to assay, or may be multi-allelic but recorded as bi-allelic in the PGM. Strand ambiguous variants can make bioinformatic analyses complicated. Multiple different polygenic models may be created to cater for different assays as a result. For example, the CanRisk tool provides a flexible platform for the integration of alternative PGMs and currently incorporates inputs for many such models into its multi-factorial breast cancer risk prediction model [4,12]. The majority of these are developed from only two underlying PGMs: PGS313 and PGS77 [12]. The set of options emerges from differing efforts to develop assays for the clinical implementation of the same original model.

PGM methods optimized for clinical utility should therefore take these considerations into account in the model development phase to facilitate consistent and straightforward implementation. In this paper, we present a refinement of the previously published S4 (select and shrink with summary statistics) PGM method that is designed to optimize performance when restricted to a pre-specified number of available SNPs while limiting candidate variants to those on chip genotyping arrays that pass stringent quality control (QC) and are likely to be assayable using other methods. We compare the performance of this restricted S4 method at predicting breast, colorectal, ovarian, and prostate cancer risk to several PGMs currently in clinical use as well as the more computationally expensive full S4 model that includes tens to hundreds of thousands of variants. These tests aim to assess model development in settings that more realistically reflect clinical use.

## METHODS

The objective of this method is to produce a PGM that facilitates the development of a clinically deployable panel. Our approach builds on the original S4 framework with modifications to tailor the number and characteristics of selected variants whilst maintaining predictive performance [13–15]. Unlike the unrestricted S4 method which selects tens to hundreds of thousands of variants for inclusion in the final model, this restricted S4 method (S4-Oligo) prioritizes producing a PGM with a smaller number of SNPs that is better suited for realistic clinical use. Limiting the variant set to well- behaved SNPs with demonstrated consistent genotyping performance reduces the risk of assay failure and simplifies the manufacturing and validation of a fixed genotyping panel. To achieve this all candidate variants were required to be non-strand- ambiguous, not an indel, and to have been successfully genotyped using the OncoArray genotyping array [16]. The rationale for this is based on the experience of clinical laboratories in the implementation of the published 313 variant breast cancer polygenic risk model for application in CanRisk [4]. Of the 313 variants, 113 were included on either the iCOGS or OncoArray genotyping arrays and passed genotyping QC [17]. For example, among five sequencing-based clinical assays developed for inclusion in the CanRisk model, between 4 to 80 of the 313 variants failed sequencing assay design [4]. Importantly, all 113 variants that were successfully genotyped were retained across the assays.

Summary statistic sources for model development varied by phenotype. Ovarian cancer summary statistics were drawn from a recent multi-ancestry GWAS [18]. These include genome-wide association results for non-mucinous ovarian cancer subtypes and were generated through a large collaborative meta-analysis combining multiple international cohorts. The summary statistics provided effect size estimates, standard errors, allele frequencies, and variant-level quality metrics across the autosomes. Breast and colorectal cancer summary statistics were identical to those used in previously published S4 model development and testing [19]. These were generated from large-scale, multi-cohort meta-analyses incorporating data from population-based biobanks and case-control studies. Only variants passing the original study’s QC filters were used in the present analysis. Summary statistics for prostate cancer were derived from a recent meta-analysis which included data from FinnGen R5, UK Biobank, and Biobank Japan along with other cohorts [20]. Because Biobank Japan contributed to the prostate meta-analysis, it could not be used as an independent test set for downstream evaluation.

All model parameters were tuned on the FinnGen dataset [21]. To increase the sample size of development data and improve accuracy, we incorporated data from the test datasets into the input summary statistics. Although no separate tuning set was available for these augmented statistics, we reused the parameters optimized on FinnGen. This approach is justified because FinnGen and Biobank Japan contributed a substantial proportion of additional data to increase precision, and the S4 parameters remain valid under variance reduction. Thus, the underlying model structure remained consistent after augmentation. This approach resembles PGM methods that use all available data for model construction, with a portion of the data implicitly serving as a tuning component through its contribution to the augmented summary statistics [22]. To ensure independence between the tuning dataset and the summary statistics for prostate cancer, the contribution of FinnGen R5 was removed prior to model development. This was done by reverse meta-analysis, algebraically subtracting the FinnGen R5 effect estimates and variances from the combined statistics to reconstruct the estimates from the non-FinnGen component to ensure independence. Additional details on this calculation are available in the Supplementary Methods.

### S4-Oligo method summary

Variants are selected by an iterative grouping and filtering process to produce an optimally informative set of SNPs (Figure 1). Variants are first ranked by ascending p- value, ensuring the most statistically significantly associated variants are processed first. Top-ranked SNPs are either added to an existing group of variants, selected as the lead variant for a new group, or discarded if it contributes redundant information with the current pool of variants. First, any existing group whose lead variant lies within 3 cM (centimorgans) of the current SNP is identified. The candidate variant is discarded if its maximum absolute correlation (*r*-squared) across these groups exceeds 0.7. This threshold was adjusted downward from the original S4 method (0.85) to balance redundancy reduction with the retention of potentially informative variants. Otherwise, the SNP is added to the group contains a variant with the highest absolute correlation. If no such group exists, a new group is formed with that candidate as the lead variant. The joint effects of adding the candidate variant to an existing group are assessed, and if the combined estimates fall outside reasonable bounds, the variant is discarded, thereby avoiding extreme positive and negative values. The number of variants selected in the final model is controlled by adjusting the p-value threshold to achieve a pre-specified number of selected variants. Models were generated with 200, 300, 500, or 1,000 variants.

**Figure 1.**
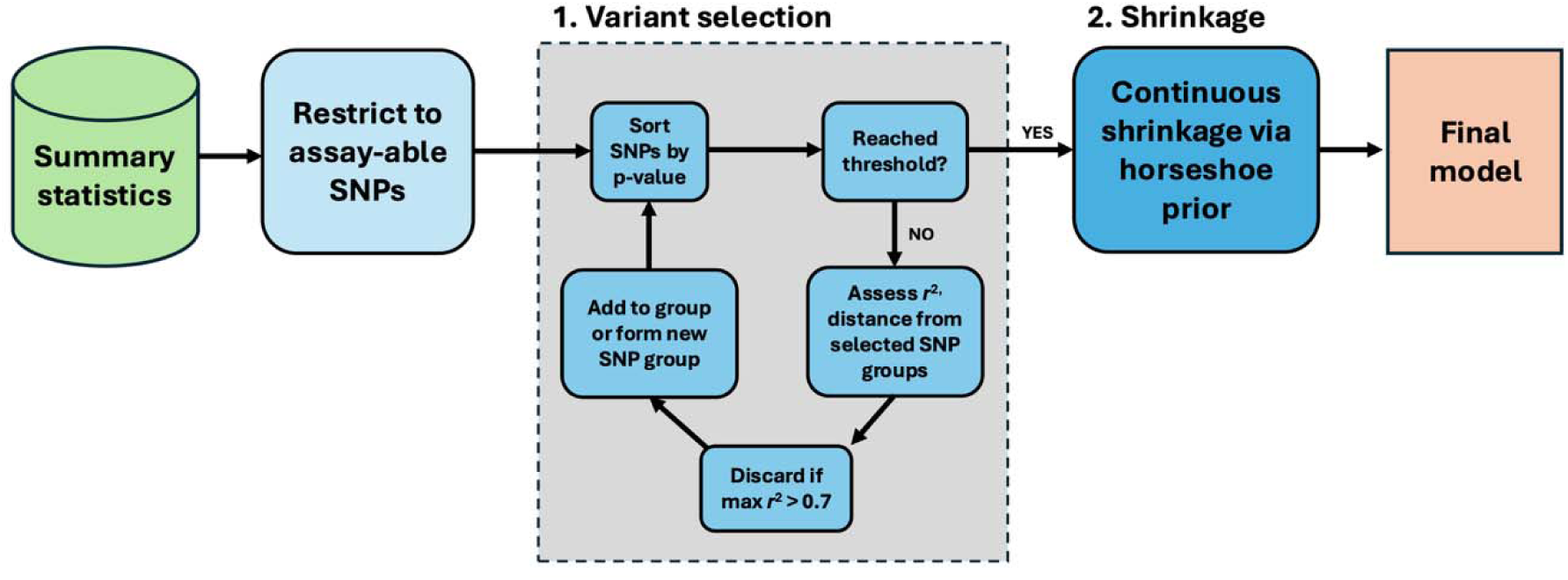
S4-Oligo model development flowchart.

Effect size calculation is then performed in the same way as the original S4 method, employing continuous shrinkage on effect sizes via a generalized horseshoe prior that suppresses noise from weaker signals while allowing strong associations to persist. The model is tuned by performing a grid search over the global and local shrinkage parameters as previously described [14]. Linkage disequilibrium (LD) is accounted for using a multi-ethnic reference panel (1000 Genomes) to enable robust modelling across diverse populations, consistent with the original S4 method [23].

### Model validation

The complete set of PGMs tested for each cancer are presented in Table 1. Published polygenic scores for each cancer were included as a benchmark, as well as the full unrestricted version of the S4 method. This enables comparison of the performance of the restricted S4 method against comparable examples, as well as with a much larger model to assess the extent to which predictive information is omitted by restricting to fewer variants.

**Table 1.** Full set of PGMs tested for each phenotype.

| Phenotype | PGM Name | Number of SNPs | Method/Source 193 |
|---|---|---|---|
| Breast cancer | Breast309 | 309 | Published PGS [7] |
|  | S4-200 | 200 | S4-Oligo |
|  | S4-300 | 300 |  |
|  | S4-500 | 500 |  |
|  | S4-1000 | 1,000 |  |
|  | S4 | 390,371 | Original S4 |
| Colorectal cancer | Colorectal103 | 103 | Published PGS [24] |
|  | S4-200 | 200 | S4-Oligo |
|  | S4-300 | 300 |  |
|  | S4-500 | 500 |  |
|  | S4-1000 | 1,000 |  |
|  | S4 | 285,405 | Original S4 |
| Ovarian cancer | Ovarian36 | 36 | Published PGS [13] |
|  | S4-200 | 200 | S4-Oligo |
|  | S4-300 | 300 |  |
|  | S4-500 | 500 |  |
|  | S4-1000 | 1,000 |  |
|  | S4 | 64,518 | Original S4 |
| Prostate cancer | Prostate269 | 269 | Published PGS [25] |
|  | Prostate451 | 451 | Published PGS [26] |
|  | S4-200 | 200 | S4-Oligo |
|  | S4-300 | 300 |  |
|  | S4-500 | 500 |  |
|  | S4-1000 | 1,000 |  |
|  | S4 | 409,227 | Original S4 |

Model testing for risk prediction in the general population was performed in UK Biobank, Biobank Japan, and the *All of Us* Research Program. Effect size estimates were obtained by fitting a logistic regression of binary disease endpoints on the standardized PGS, adjusting for the first four genetic principal components and age at diagnosis for cases or age at last follow up for controls. Case and control counts by phenotype and biobank are provided in Table S1. Phenotypes in UK Biobank were derived using the ukbrapR R package to extract disease endpoints [27]. Ovarian cancer cases were restricted to non-mucinous histology based on available ICD-O-3 histology codes. All eligible cases were included to maximize statistical power, since excluding subtypes from the relatively small ovarian cancer case set would have limited the ability to detect meaningful differences between models. Because breast and colorectal cancers had sufficiently large case numbers, only incident cases were retained to minimize population stratification effects and reduce survival-related confounding while retaining statistical power. The summary statistics used in prostate cancer model development incorporated UK Biobank data published in early 2023. To account for this, only cases diagnosed after January 1, 2022 were included, while earlier cases were excluded. Restricting to post-2022 incident cases ensured that the UK Biobank test data remained independent of the training summary statistics. Disease status in *All of Us* was determined by applying electronic health record-based algorithms for ovarian, breast, and prostate cancers. Prior work suggests that a high sensitivity criteria that includes all participants with any occurrence of qualifying condition code as cases is likely to maximize power for colorectal cancer association analysis [28]. We adopt that case definition here.

Studies from the Consortium of Investigators of Modifiers of *BRCA1*/*2* (CIMBA) have demonstrated that PGM developed for breast, ovarian, and prostate cancer prediction in the general population also modify risks of those cancers for carriers of pathogenic variants in both *BRCA1* and *BRCA2* genes (*BRCA1/2*) [13,29–31]. We also test the performance of restricted S4 models (200, 300, 500, 1,000 SNPs) against the same set of published PGSs in *BRCA1/2* pathogenic variant carriers using the previously published methodology [31–35].

To contextualize the practical impacts of these observed differences in population level accuracy, we also considered the net reclassification of individuals into disease- specific risk strata under different PGMs. Risk groups can be defined by thresholds for clinical action. The goal is to assess the number of individuals who would receive different clinical recommendations based on the estimated lifetime risk under each PGM. We categorized risk groups for breast cancer based on NICE guidelines for clinical action, using lifetime risk thresholds of 17% and 30% to split participants into three groups: near-population risk (< 17%), moderate risk (17-30%) and high risk (> 30%) [36]. Similarly, for ovarian cancer we used lifetime risk thresholds of 5% and 10% to define the same three groups. We performed this stratification using three different PGMs: a published PGM for each disease, the S4-200 model, and the S4-1000 model. Individuals consisted of all women from the UK Biobank cohort to provide a large sample to observe the classification differences between models. Population risk estimates were obtained from national incidence statistics and modeled using cubic splines, with lifetime risk estimated by combining this baseline risk with the relative risk from the PGS for each individual. Similarly, we also simulated risk stratification for *BRCA1/2* pathogenic variant carriers by applying gene-specific relative risks to this same cohort.

## RESULTS

Overall results (odds ratios per PGS standard deviation) for prediction in the general population are presented in Figure 2. In subsequent discussion and performance comparison, percent differences in performance are expressed on the log scale (as log odds ratios) to avoid understating differences at small effect sizes. Complete results stratified by all combinations of training data, PGM method, model size, and validation data in this study are provided in Table S2 along with additional performance metrics of area under the ROC curve (AUC) and log likelihood chi-square statistics where available.

**Figure 2.**
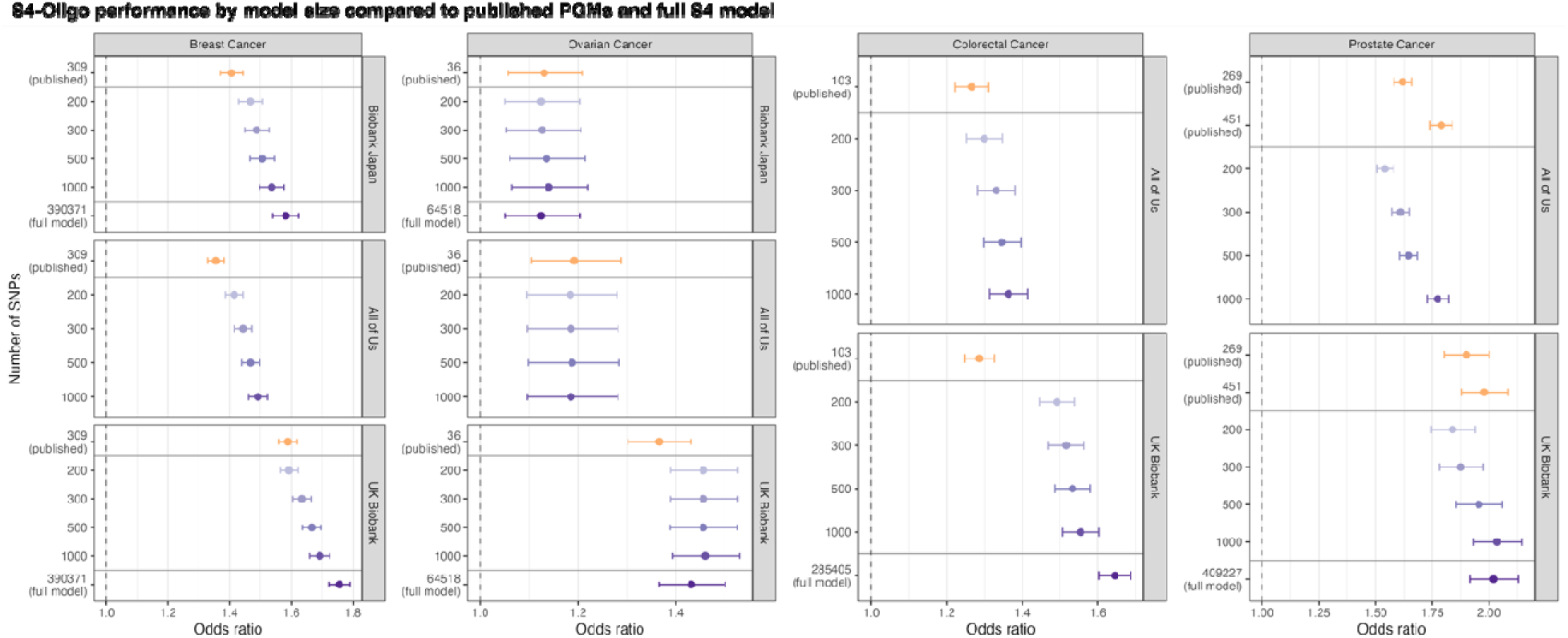
Restricted S4 model performance (odds ratio per standard deviation) by number of SNP: Published PGS (orange) and full S4 model included for comparison. The full S4 model was not run in *All of Us* due to cost constraints. 95%-CI are shown.

Restricted S4 models outperformed their published counterparts with the most similar number of variants at breast and colorectal cancer prediction. The restricted S4 model closest in size to its published counterpart achieved 21% improvement for breast cancer prediction (S4-300 vs. Breast309) and 52% improvement for colorectal cancer prediction (S4-200 vs. Colorectal103). The S4-1000 models further improved prediction accuracy over published PGMs, achieving 28% and 69% improvement for breast and colorectal cancers, respectively. For prostate cancer prediction, the published PGMs tended to outperform the restricted S4 methods at a given model size. For example, Prostate451 outperformed S4-500 but fell short of the accuracy of S4-1000. This behavior was consistent in both *All of Us* and UK Biobank. For ovarian cancer prediction, accuracy varied considerably by biobank. While all S4 models outperformed Ovarian36 in UK Biobank, we observed limited differences between models in *All of Us* and Biobank Japan in general.

In general, the marginal gain in predictive accuracy from additional SNPs diminished with model size. While the full S4 model generally achieved top performance except for ovarian cancer prediction, the S4-1000 model was typically closer in performance to the full S4 model than the published PGS while including between 0.2% and 2% of the number of variants. This suggests that most of the predictive information leveraged by the full S4 method is contained in a relatively small number of variants with diminishing returns from increased model size. Additionally, all models showed reduced performance in the A*ll of Us* cohort. This is expected given the known challenges of PGS portability in ancestrally diverse populations [37–39]. Results stratified by ancestry suggest that this performance gap was primary driven by reduced performance in African-origin ancestries (Figure S1). While this was not the case for ovarian cancer, wide confidence bounds limit comparisons between performance in *All of Us* and Biobank Japan.

Results in for risk prediction in carriers of pathogenic *BRCA1* or *BRCA2* (*BRCA1/2*) variants are presented in Figure 3. We observed attenuated performance in *BRCA1/2* carriers relative to the general population for both breast and prostate cancer prediction. While there is greater uncertainty in these estimates due to more limited sample sizes, the overall performance trends mirror the results in the general population. Though the larger models show performance improvement, most predictive information contained in a relatively small subset of SNPs with diminishing returns from including additional variants.

**Figure 3.**
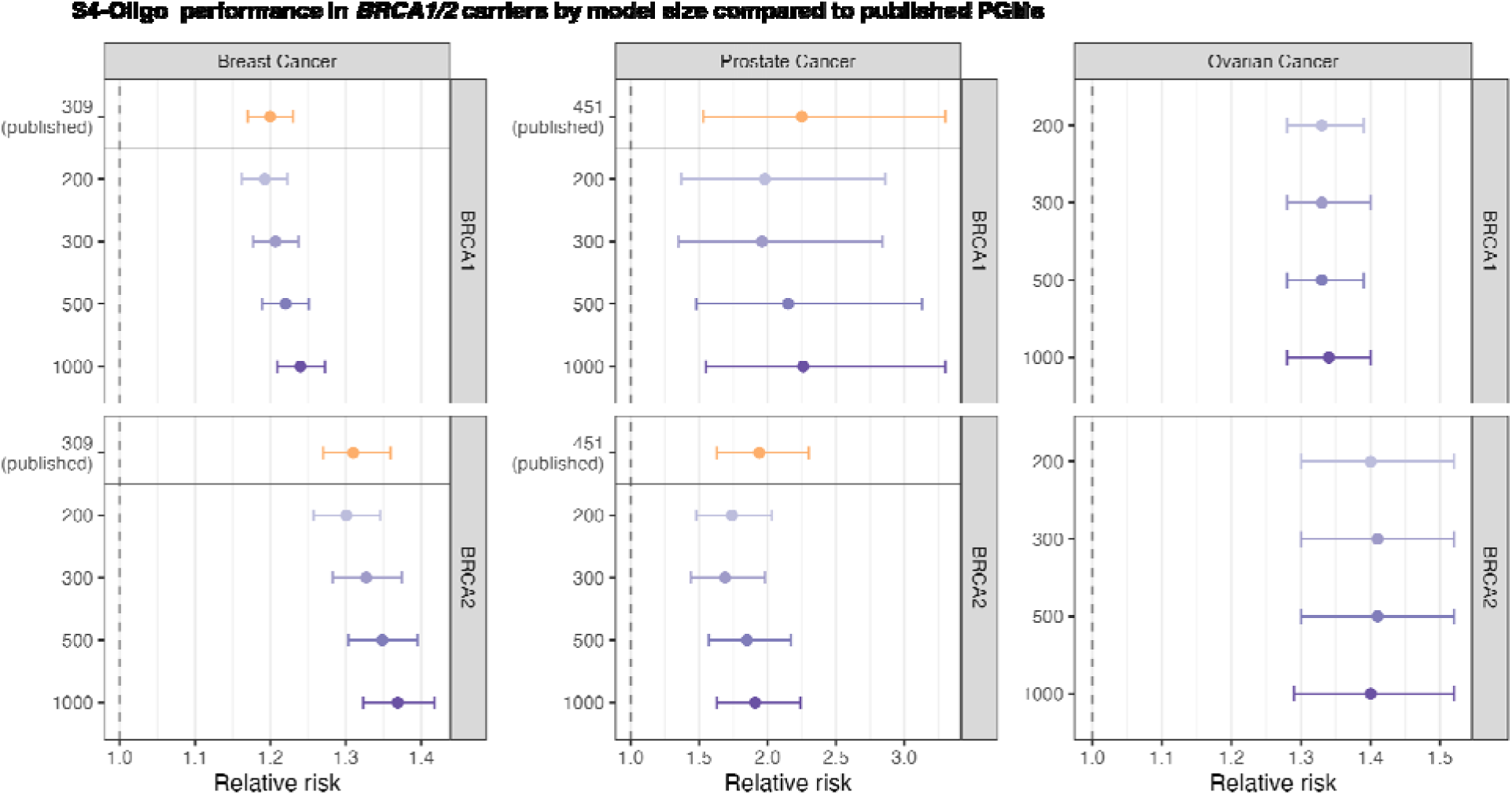
S4-Oligo performance in carriers of pathogenic *BRCA1* and *BRCA2* variants: 95%-CI are shown. Relative risks are hazard ratios for breast cancer models, and odds ratios for prostate and ovarian cancer models.

Net reclassification results for breast and ovarian cancer into risk strata under S4-200, S4-1000, and the full S4 model are presented in Figure 4. The biggest changes under the more accurate models came in breast cancer prediction. This is both because of the strong performance of S4-Oligo relative to the published PGM in breast cancer prediction as well as the higher baseline risk in the population putting more individuals closer to the relevant decision-making thresholds for lifetime risk. For both *BRCA1 and BRCA2* carriers, the net reclassification from larger models came from reclassifying higher risk individuals into lower risk groups. This was most pronounced for *BRCA2* carriers and ovarian cancer risk. Incorporating PGS into risk prediction resulted in more reclassification among *BRCA2* carriers than *BRCA1* carriers for both breast and ovarian cancer. This is to be expected by the lower penetrance of *BRCA2* relative to *BRCA1* placing a greater number of participants closer to the relevant risk thresholds for clinical action. Nearly all individuals with *BRCA1* variants are predicted to have high lifetime risk, so the PGSs add little additional predicative value. However, among *BRCA2* carriers, 7% of individuals classified as high risk are re-classified into a lower risk group by adopting the smallest S4-Oligo model.

**Figure 4.**
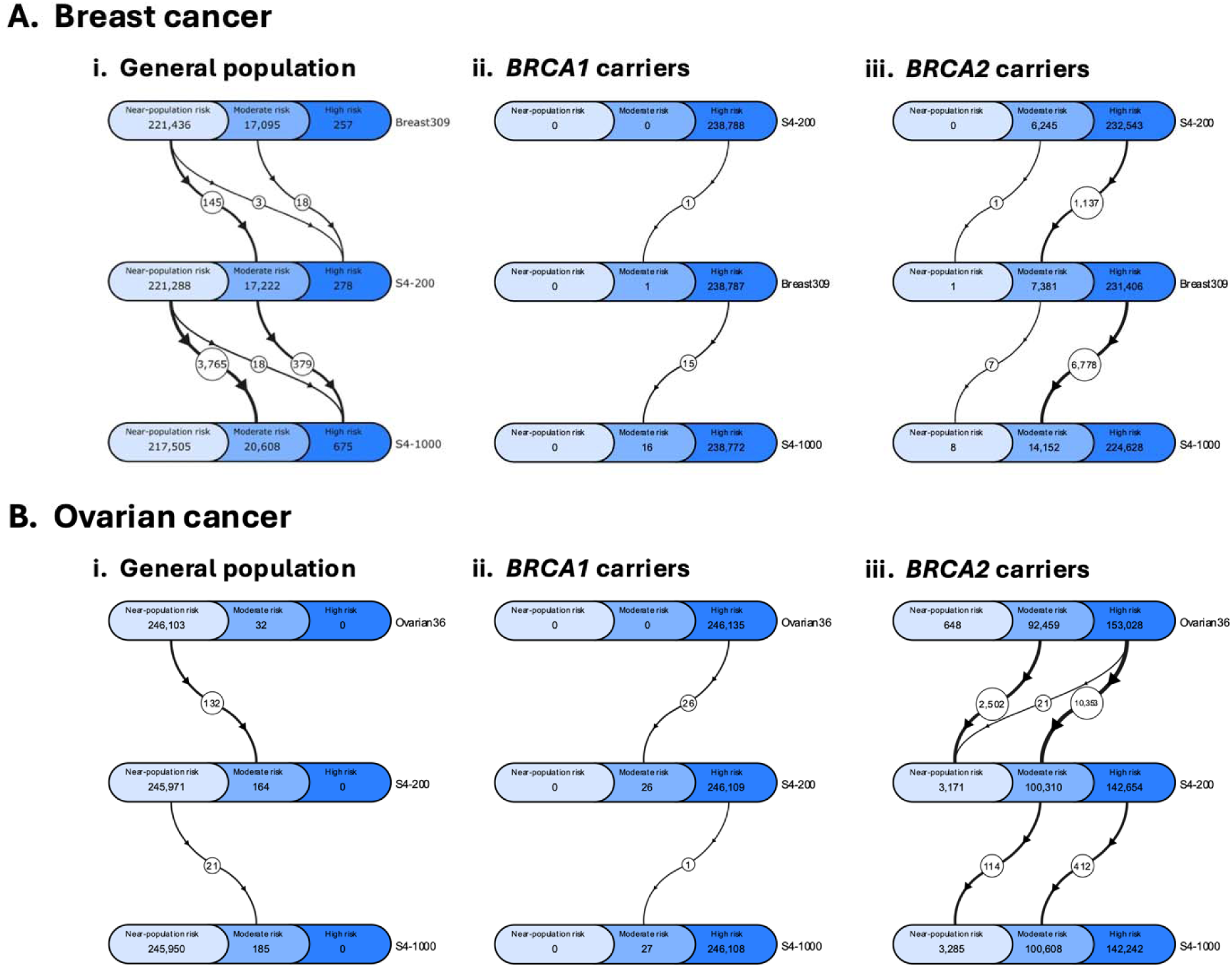
Net change in disease risk classification under different PGMs: Risk groups are defined based on estimated lifetime risk and NICE clinical guidelines. In all panels, the pool of individuals consists of women from UK Biobank. For breast cancer, near-population risk (< 17%), moderate risk (17- 30%), and high risk (> 30%). For ovarian cancer, near-population risk (< 5%), moderate risk (5-10%), and high risk (> 10%). PGMs are ordered from least accurate (top) to most accurate (bottom). Circled values indicate the net flow of participants between the linked groups. Full 3×3 contingency tables showing both inflow and outflow are available in Table S3.

## DISCUSSION

In this study, we presented and evaluated S4-Oligo, a PGM method developed to produce models that are well-suited for realistic clinical implementation by restricting model size and candidate SNPs. We demonstrated that this approach retains much of the predictive power of larger unrestricted models while matching or exceeding the performance of published PGMs developed with clumping and thresholding for breast, colorectal, prostate, and ovarian cancer risk prediction in *All of Us*, UK Biobank, and Biobank Japan, as well as breast and prostate cancer risk-modification in *BRCA1/2* carriers in CIMBA. To date, most work developing PGM methods by utilizing penalized regression or machine learning approaches has focused on optimizing models to maximize accuracy irrespective of model size. Our results illustrate the degree to which most of the predictive information from these large PGMs is contained in a relatively small number of SNPs with diminishing returns as model size increases. While the restricted models fell short of the performance of the full S4 model as expected, the gap was small relative to the substantial decrease in model size. The largest tested S4- Oligo method (S4-1000) retained 92% (breast cancer), 88% (colorectal cancer), 100% (prostate cancer), and 104% (ovarian cancer) of the performance in log odds ratio of the full S4 method despite including only between 0.2% and 2% of the number of SNPs. Even the smallest 200 SNP model retained 84%, 80%, 101%, and 86% of performance, respectively, relative to the full S4 model. While much larger models did tend to offer performance gains, this comes at the tradeoff of clinical portability.

We found no evidence that S4-Oligo offered improvement over size equivalent published PGMs for prostate and ovarian cancer prediction. For ovarian cancer, there was little evidence of differences between models in general, including the full S4 model. Though relatively small case set sizes produced estimates with a large degree of uncertainty, this trend was observed across all three biobanks, so this may also reflect the less polygenic architecture of ovarian cancer risk. Ovarian cancer performance was considerably lower overall relative to the other disease endpoints. Prostate cancer models were more varied in their performance with increasing model size. While S4-300 and S4-500 matched but did not exceed the performance of Prostate269 and Prostate451, respectively, the published models were based on a larger training data set. These results demonstrate that computationally intensive methods can achieve performance gains over published PGMs when tailored to produce models of comparable size. The improvements at breast and colorectal cancer risk prediction suggest that the S4-Oligo method successfully leveraged limited genetic information by optimizing SNP selection and weighting more efficiently than published PGMs in these settings. We encourage future work to apply these methodological advances to better-maximize the information retained in a limited set of variants and optimize the performance tradeoff relative to the unrestricted model. Notably, while these advances in PGM methodology have made meaningful gains in accuracy, the practical differences between models remain relatively small at the population level, as highlighted by Figure 4.

We observed attenuated performance across all models in carriers of *BRCA1* and *BRCA2* variants for breast and prostate cancer (Figure 3). This is consistent with prior work on PGS modification of risk among individuals with these variants [31]. The predicted cumulative lifetime risk of breast cancer in the general population as well as in *BRCA1/2* carriers under these same PGMs is shown in Figure S2. While the effect sizes of all PGMs are attenuated in carriers, the net reclassification grouped by clinically actionable thresholds is more substantial due to those individuals having a higher baseline risk while also reaching risk-based clinical action thresholds at earlier life stages (Figure S2). Together, these results highlight how the impact a PGS on risk stratification involves an interaction between the baseline disease risk in the general population, the effect size of the PGM, and disease-specific clinical decision-making thresholds. For instance, the net impact of including the PGS in ovarian cancer risk stratification was much greater in *BRCA2* carriers than in the general population despite a lower relative risk of each PGS within each group. This is because of the low baseline risk for ovarian cancer in the general population and relatively high threshold for clinical action from identifying an individual as having high lifetime ovarian cancer risk.

The lack of portability of PGSs to non-European-origin ancestries is well- documented and remains a central obstacle to equitable PGS implementation [38,39]. Results in *All of Us* stratified by genetically inferred ancestry group are presented in Table S1. As expected, performance tends to decrease as the genetic distance from the input summary statistic population grows, with especially notable declines in accuracy for prediction in African-origin ancestries. This was the primary driver of the reduced performance in *All of Us* relative to UK Biobank and Biobank Japan. Additionally, PGS distributions are known to vary by country even within European- origin ancestries [40]. This suggests that optimizing accuracy for clinical use likely means taking a both an individual’s genetic ancestry and country of origin into account to better convert their PGS into a risk estimate as previously suggested [41].

In this analysis, S4-Oligo was developed from a pool of SNPs selected because they are expected to be well-suited to developing clinical assays. In principle, any additional selection criteria could be imposed to this candidate set before model fitting and tuning depending on downstream limitations. The approach here outlined serves as a generalizable framework that can be adapted to specific use cases. However, evidence suggests that the specific criteria presented in this analysis represents a reasonable evaluation of model performance under the requirement of assay-able SNPs.

## CONCLUSIONS

The method presented here demonstrates how computationally intensive PGM methods can be leveraged to optimized PGM development for clinical use. Models limited to fewer than 1,000 SNPs were able to match or exceed published PGMs for several cancers which retained most of the predictive performance of models containing tens or hundreds of times the number of SNPs. Tailoring PGM methods for sizes and contexts where PGSs can provide optimal stratification value such as modifying risk for carriers of high-penetrance risk alleles should be a central focus moving forward.

## Data Availability

Data used in this study are available from the All of Us Research Program (https://www.joinallofus.org/), Biobank Japan (https://biobankjp.org/), FinnGen (https://www.finngen.fi/), UK Biobank (https://www.ukbiobank.ac.uk/), and CIMBA (https://www.ccge.medschl.cam.ac.uk/consortium-investigators-modifiers-brca12-cimba). Code used in this study is available upon request to the authors.

## ACKNOWLEDGEMENTS

We thank *All of Us* participants for their contributions. We also thank the *All of Us* Research Program for making available the participant data examined in this study. We affirm that the procedures followed in this analysis were in accordance with ethical standards on human experimentation and that proper informed consent was obtained. This research has been conducted using the UK Biobank Resource under Application Number 28126.

## SUPPLEMENTAL TABLES

**Table S1.** Case counts by biobank and phenotype: Note that UK Biobank counts are estimated from standard errors of effect sizes due to the ongoing shutdown

| **Phenotype** | **Study** | **Cases** | **Controls** |
|---|---|---|---|
| Breast cancer | Biobank Japan | 6,325 | 73,225 |
|  | FinnGen | 24,270 | 222,078 |
|  | UK Biobank | 11,275 | 200,000 |
| Colorectal cancer | Biobank Japan | 8,305 | 159,386 |
|  | FinnGen | 11,790 | 378,749 |
|  | UK Biobank | 4,046 | 400,000 |
| Ovarian cancer | Biobank Japan | 843 | 60,614 |
|  | FinnGen | 2,339 | 222,078 |
|  | UK Biobank | 1,735 | 200,000 |
| Prostate cancer | Biobank Japan | 5,672 | 84,660 |
|  | FinnGen | 20,368 | 156,671 |
|  | UK Biobank | 1,410 | 200,000 |
| Breast cancer – *BRCA1* carriers | CIMBA | 9,240 | 10,533 |
| Breast cancer – *BRCA2* carriers | CIMBA | 6,345 | 6,994 |
| Prostate cancer – *BRCA1* carriers | CIMBA | 70 | 365 |
| Prostate cancer – *BRCA2* carriers | CIMBA | 130 | 880 |

**Table S2.**
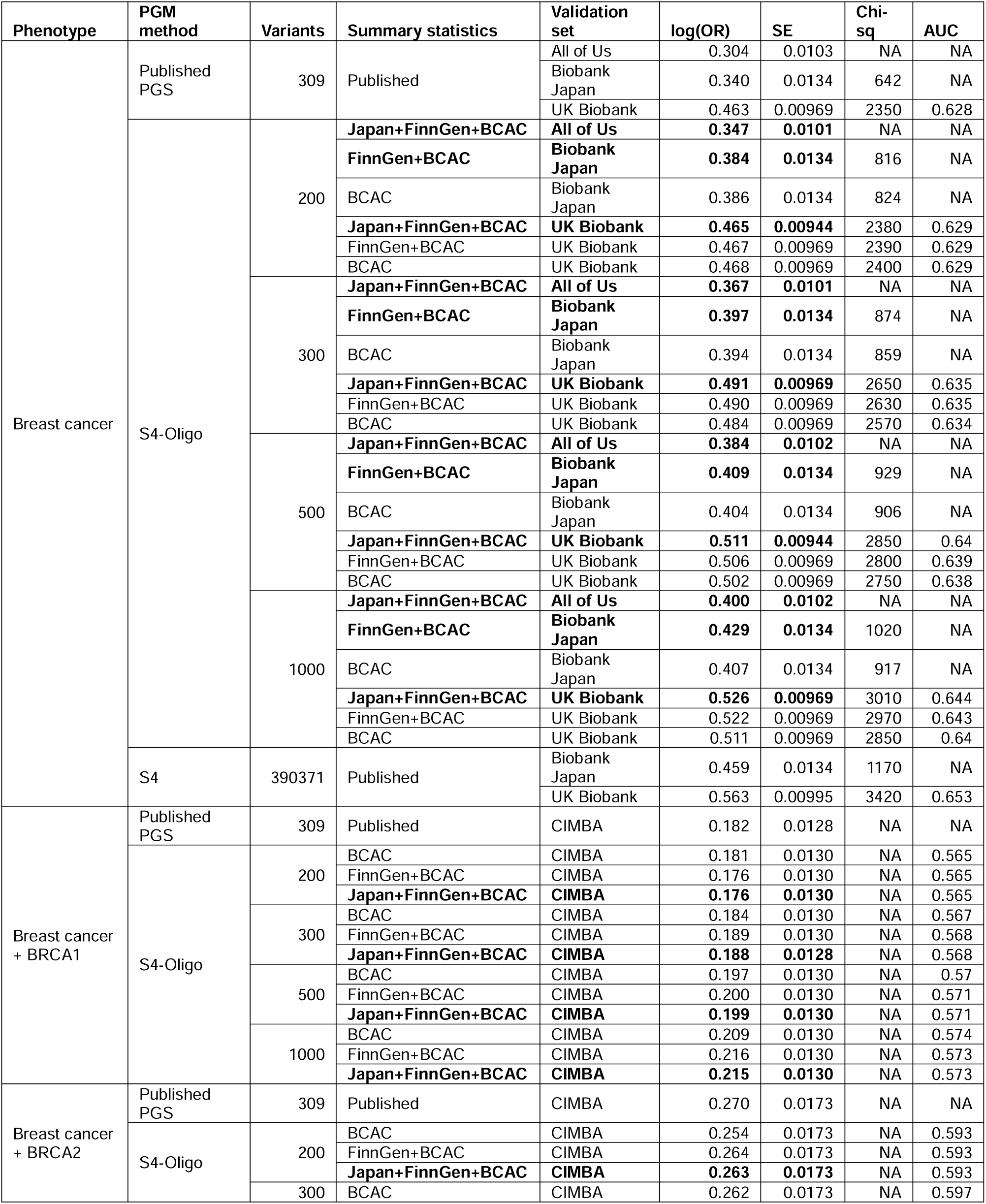
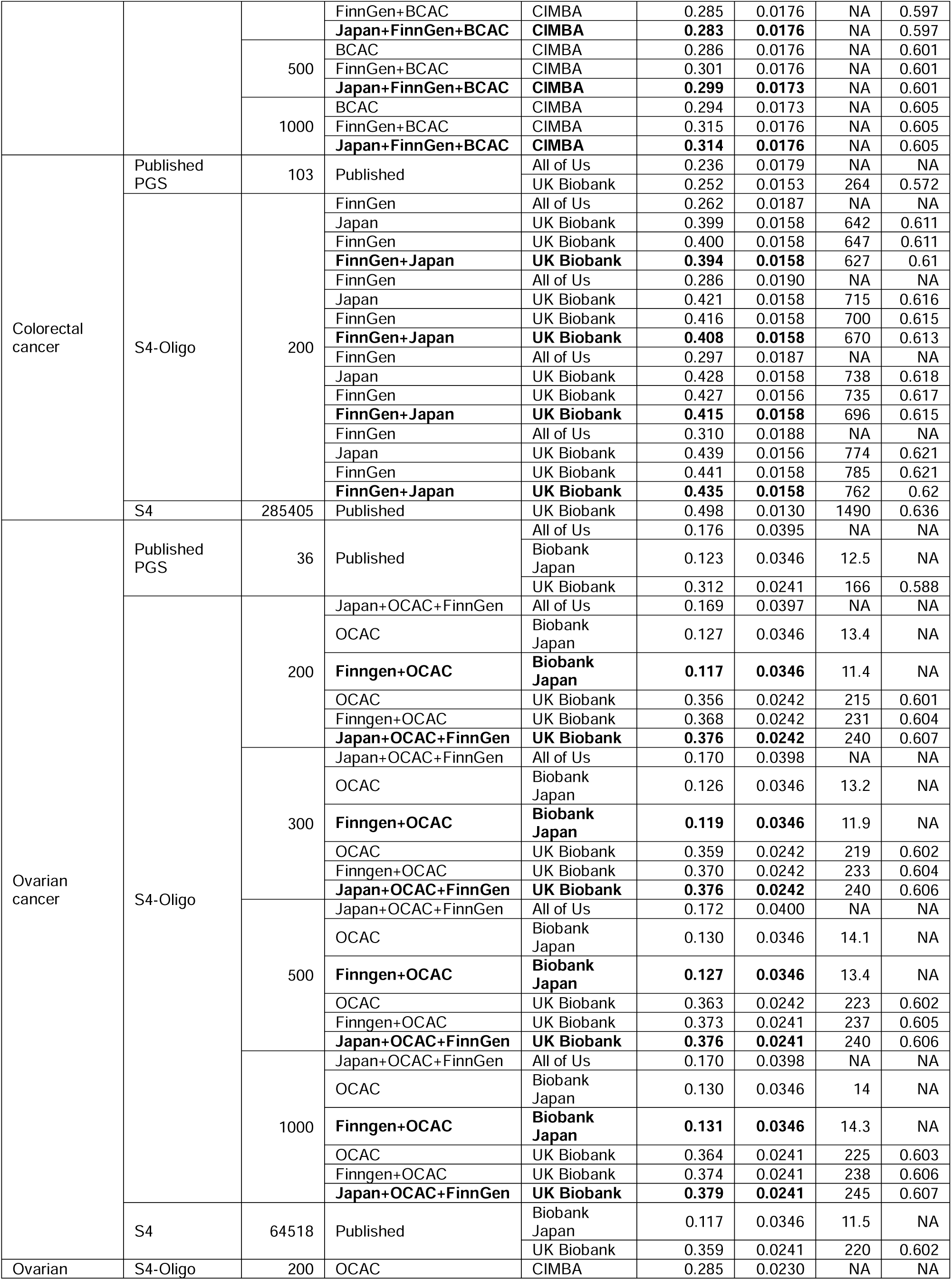
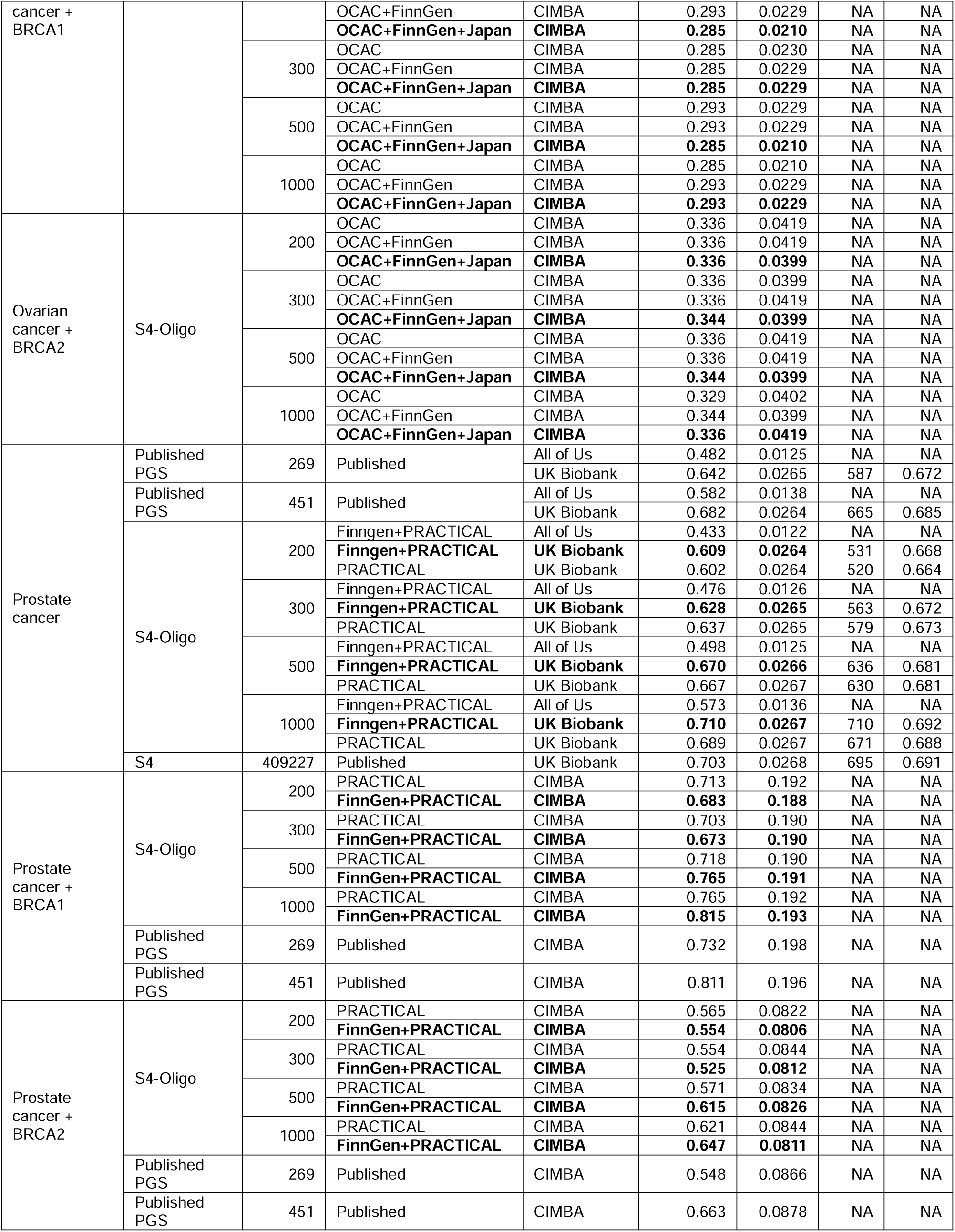
Full results table, all performance metrics: Note that not all performance metrics were computed for all runs. Values for log(OR) are scaled per standard deviation of the PGS. Summary statistic sources for the unrestricted S4 method were identical to those presented in Baierl et al 2025. The S4-Oligo models plotted in the main results table (Figure 2) are bolded.

**Table S3.** Total reclassification contingency tables: For breast cancer, risk groups are defined based on lifetime risk as follows: near-population risk (< 17%), moderate risk (17-30%), and high risk (> 30%). For ovarian cancer, risk groups are defined as follows: near-population risk (< 5%), moderate risk (5- 10%), and high risk (> 10%).

| Phenotype | PGM 1 | PGM 2 | PGM 1 Group | PGM 2 Group |  |  |
|---|---|---|---|---|---|---|
|  |  |  |  | Near-population risk | Moderate risk | High risk |
|---|---|---|---|---|---|---|
| Breast cancer | PGS313 | S4-200 | Near-population risk | 213141 | 8284 | 11 |
|  |  |  | Moderate risk | 8139 | 8761 | 195 |
|  |  |  | High risk | 8 | 177 | 72 |
|  | PGS313 | S4-1000 | Near-population risk | 209874 | 11497 | 65 |
|  |  |  | Moderate risk | 7620 | 8970 | 505 |
|  |  |  | High risk | 11 | 141 | 105 |
|  | S4-200 | S4-1000 | Near-population risk | 211634 | 9634 | 20 |
|  |  |  | Moderate risk | 5869 | 10861 | 492 |
|  |  |  | High risk | 2 | 113 | 163 |
| Breast cancer + *BRCA1* | PGS313 | S4-200 | Moderate risk | 0 | 0 | 1 |
|  |  |  | High risk | 0 | 0 | 238787 |
|  | PGS313 | S4-1000 | Moderate risk | 0 | 0 | 1 |
|  |  |  | High risk | 0 | 16 | 238771 |
|  | S4-200 | S4-1000 | High risk | 0 | 16 | 238772 |
| Breast cancer + *BRCA2* | PGS313 | S4-200 | Near-population risk | 0 | 1 | 0 |
|  |  |  | Moderate risk | 0 | 2873 | 4508 |
|  |  |  | High risk | 0 | 3371 | 228035 |
|  | PGS313 | S4-1000 | Near-population risk | 0 | 1 | 0 |
|  |  |  | Moderate risk | 8 | 4191 | 3182 |
|  |  |  | High risk | 0 | 9960 | 221446 |
|  | S4-200 | S4-1000 | Moderate risk | 8 | 4653 | 1584 |
|  |  |  | High risk | 0 | 9499 | 223044 |
| Ovarian cancer | PGS36 | S4-200 | Near-population risk | 245951 | 152 | 0 |
|  |  |  | Moderate risk | 20 | 12 | 0 |
|  | PGS36 | S4-1000 | Near-population risk | 245939 | 164 | 0 |
|  |  |  | Moderate risk | 11 | 21 | 0 |
|  | S4-200 | S4-1000 | Near-population risk | 245909 | 62 | 0 |
|  |  |  | Moderate risk | 41 | 123 | 0 |
| Ovarian cancer + *BRCA1* | PGS36 | S4-200 | High risk | 0 | 26 | 246109 |
|  | PGS36 | S4-1000 | High risk | 0 | 27 | 246108 |
|  | S4-200 | S4-1000 | Moderate risk | 0 | 20 | 6 |
|  |  |  | High risk | 0 | 7 | 246102 |
| Ovarian cancer + *BRCA2* | PGS36 | S4-200 | Near-population risk | 435 | 213 | 0 |
|  |  |  | Moderate risk | 2715 | 69879 | 19865 |
|  |  |  | High risk | 21 | 30218 | 122789 |
|  | PGS36 | S4-1000 | Near-population risk | 520 | 128 | 0 |
|  |  |  | Moderate risk | 2761 | 73947 | 15751 |
|  |  |  | High risk | 4 | 26533 | 126491 |
|  | S4-200 | S4-1000 | Near-population risk | 2468 | 703 | 0 |
|  |  |  | Moderate risk | 817 | 90495 | 8998 |
|  |  |  | High risk | 0 | 9410 | 133244 |

## SUPPLEMENTAL FIGURES

**Figure S1.**
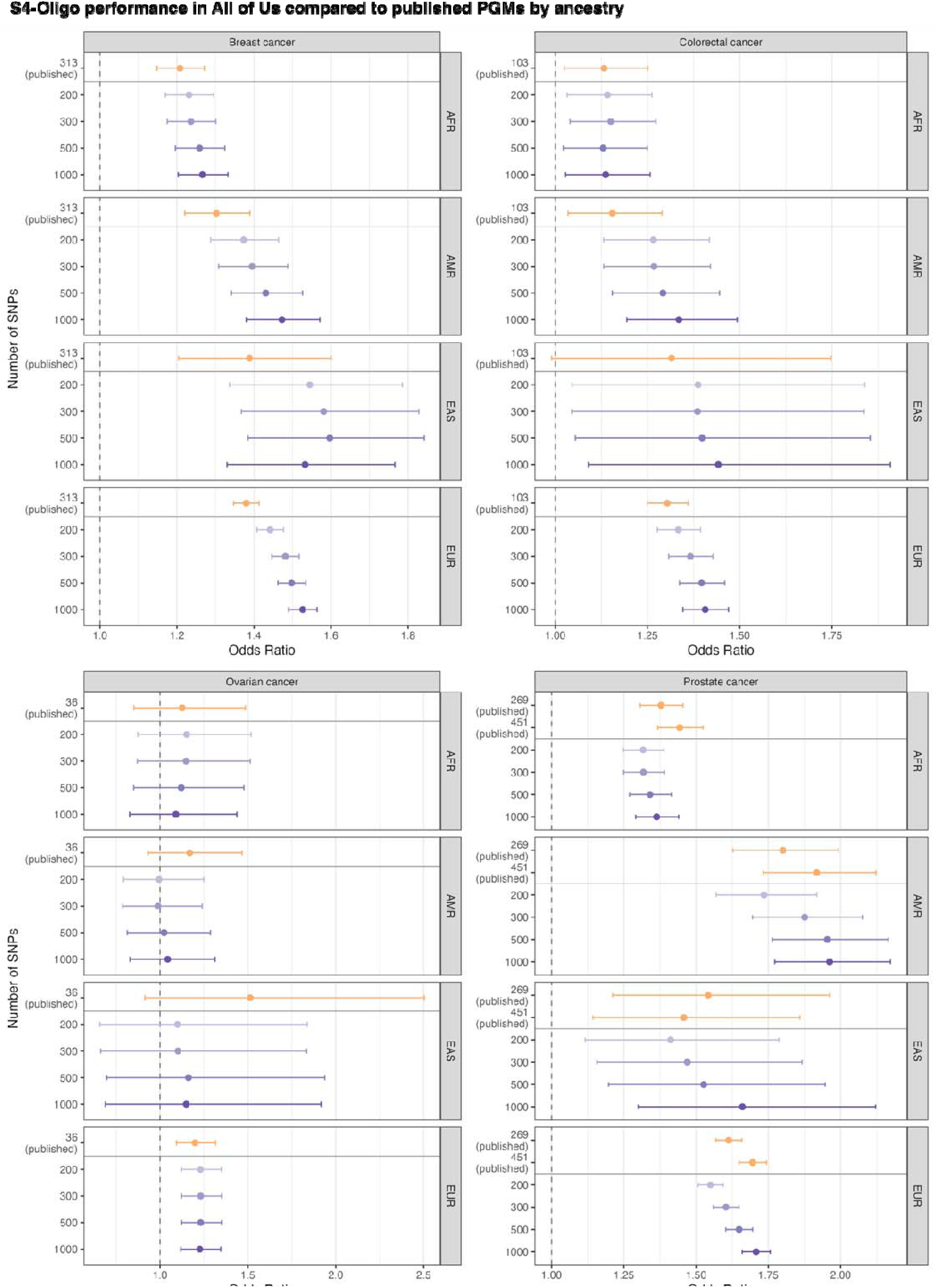
Risk prediction in *All of Us* stratified by genetically inferred ancestry: All other groups of ancestries omitted due to small case set sizes and unstable effect size estimates

**Figure S2.**
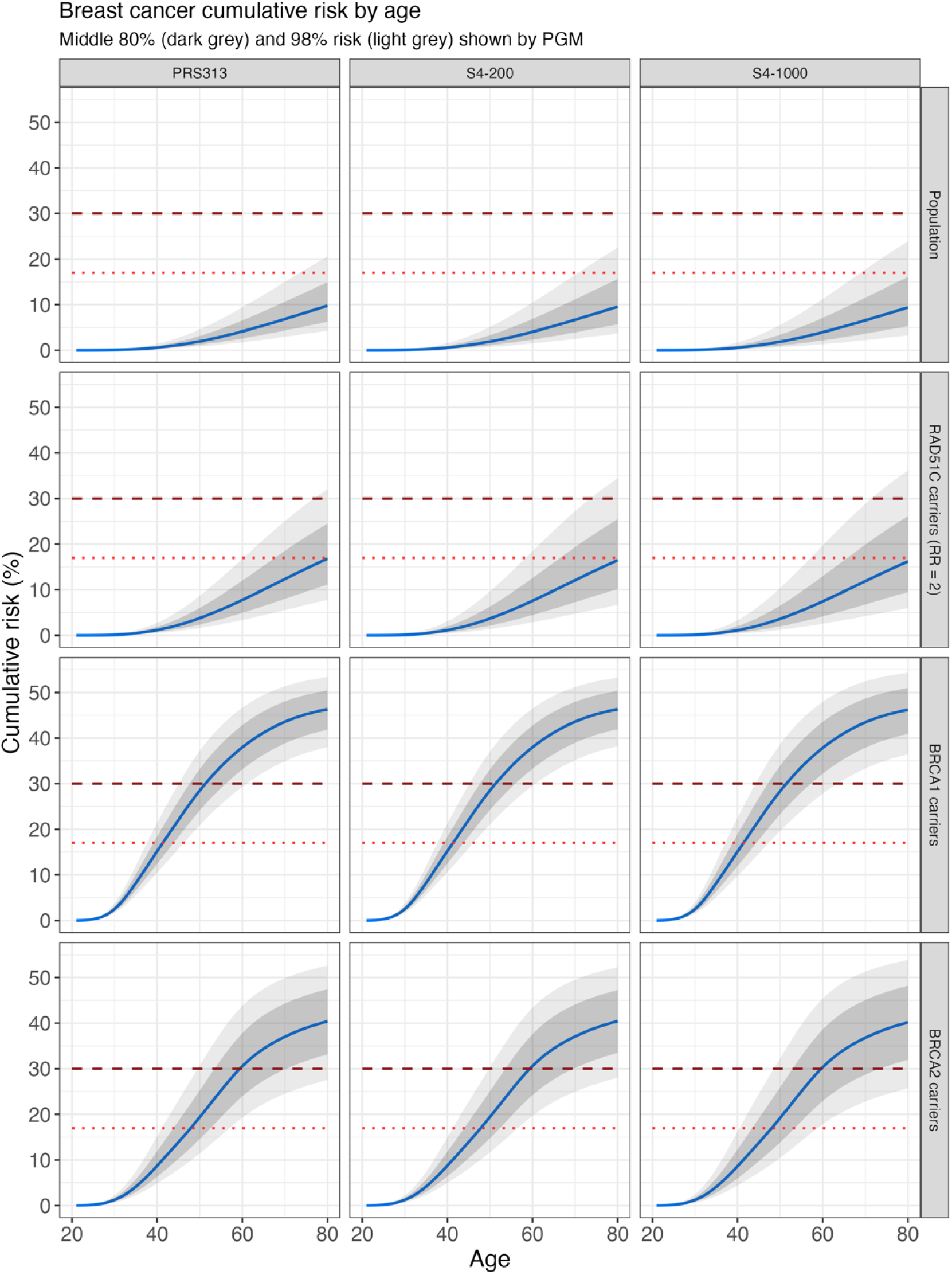
Cumulative breast cancer risk by age 80 under different PGMs: Breast cancer-specific mortality was estimated by multi-variable fractional polynomial regression of age-specific rates from national data. Age-specific penetrance of BRCA1/2 variants was computed by restricted cubic splines. RAD51D distributions use the PGS effect sizes from population prediction with a gene-specific relative risk of 2. Thresholds for medium (> 17%) and high lifetime risk (> 30%) are shown.

## SUPPLEMENTAL METHODS

Let ᶿ^ denote the meta-analyzed effect size of a particular over all combined studies with inverse variance weighting and V^ denote the pooled variance estimate. Separating the effects of FinnGen from the remaining studies, we can express these as:

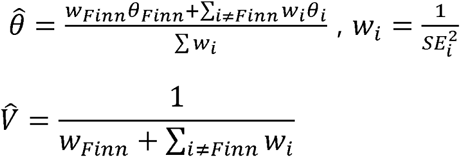

Thus, the pooled estimates after removing the influence of FinnGen can be expressed as:

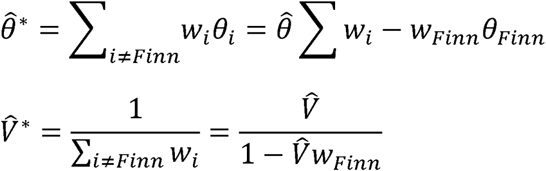

